# Quantitative CT of Normal Lung Parenchyma and Small Airways Disease Topologies are Associated With COPD Severity and Progression

**DOI:** 10.1101/2023.05.26.23290532

**Authors:** Alexander J. Bell, Ravi Pal, Wassim W. Labaki, Benjamin A. Hoff, Jennifer M. Wang, Susan Murray, Ella A. Kazerooni, Stefanie Galban, David A. Lynch, Stephen M. Humphries, Fernando J. Martinez, Charles R. Hatt, MeiLan K. Han, Sundaresh Ram, Craig J. Galban

## Abstract

**Objectives:** Small airways disease (SAD) is a major cause of airflow obstruction in COPD patients, and has been identified as a precursor to emphysema. Although the amount of SAD in the lungs can be quantified using our Parametric Response Mapping (PRM) approach, the full breadth of this readout as a measure of emphysema and COPD progression has yet to be explored. We evaluated topological features of PRM-derived normal parenchyma and SAD as surrogates of emphysema and predictors of spirometric decline.

**Materials and Methods:** PRM metrics of normal lung (PRM^Norm^) and functional SAD (PRM^fSAD^) were generated from CT scans collected as part of the COPDGene study (n=8956). Volume density (V) and Euler-Poincaré Characteristic (χ) image maps, measures of the extent and coalescence of pocket formations (i.e., topologies), respectively, were determined for both PRM^Norm^ and PRM^fSAD^. Association with COPD severity, emphysema, and spirometric measures were assessed via multivariable regression models. Readouts were evaluated as inputs for predicting FEV_1_ decline using a machine learning model.

**Results:** Multivariable cross-sectional analysis of COPD subjects showed that V and χ measures for PRM^fSAD^ and PRM^Norm^ were independently associated with the amount of emphysema. Readouts χ^fSAD^ (β of 0.106, p<0.001) and V^fSAD^ (β of 0.065, p=0.004) were also independently associated with FEV_1_% predicted. The machine learning model using PRM topologies as inputs predicted FEV_1_ decline over five years with an AUC of 0.69.

**Conclusions:** We demonstrated that V and χ of fSAD and Norm have independent value when associated with lung function and emphysema. In addition, we demonstrated that these readouts are predictive of spirometric decline when used as inputs in a ML model. Our topological PRM approach using PRM^fSAD^ and PRM^Norm^ may show promise as an early indicator of emphysema onset and COPD progression.

## Introduction

Chronic obstructive pulmonary disease (COPD) is a leading cause of death and healthcare burden in the United States and worldwide. Accounting for over 3 million deaths globally in 2015^1^, this disease is expected to rise in prevalence as the world population ages^2^. COPD is understood to be a complex heterogeneous disease presenting clinically diverse phenotypes^3, 4^. Major causes of airflow obstruction are attributed to chronic bronchiolar obstruction, a.k.a small airways disease (SAD), and emphysema. Although SAD and emphysema are treated as separate COPD subtypes, studies have shown strong quantitative evidence that SAD exists as an intermediate state between healthy lung tissue and emphysema—i.e., irreversible lung damage—in COPD pathogenesis^5–7^. At present, little has been done to better quantify the onset of SAD from healthy lung parenchyma.

The Parametric Response Map (PRM) is a CT-based voxel-wise computational technique that can identify and quantify functional small airways disease (fSAD; an indirect measure of SAD) even in the presence of emphysema^8^. The percent volume of PRM-derived fSAD (PRM^fSAD^), i.e., the amount of fSAD in the lungs, has improved COPD phenotyping and the prediction of spirometric decline in subjects at risk of COPD^9^. To determine the value of spatial features from each PRM classification, we developed topological PRM (tPRM) as an extension of the PRM algorithm^10^. These radiographic tPRM readouts were shown to improve upon commonly used whole-lung PRM measures with respect to COPD characterization, and correlate to structural changes in lung tissue samples from lung transplant recipients diagnosed with bronchiolitis obliterans^11^.

In this study, we evaluated the PRM topologies volume density (V), a measure of extent, and Euler-Poincaré Characteristic (χ), a measure of pocket formation, of normal lung and fSAD as independent readouts of COPD severity, pulmonary function, and extent of emphysema using the Phase 1 COPDGene cohort^12^. We also investigated the potential of these topologic readouts as predictors of spirometric decline using a machine-learning model. This study demonstrates how tPRM readouts may be used as possible measures of early emphysema and COPD progression.

## Materials and Methods

### Study Sample

Our study was a secondary analysis of data from COPDGene (ClinicalTrials.gov: NCT00608764), a large Health Insurance Portability and Accountability Act-compliant prospective multi-center observational study. In Phase 1 (2007-2012) and Phase 2 (2013-2017), 5-year follow-up, written and informed consent was obtained from all participants and the study was approved by local institutional review boards of all 21 centers. Ever-smokers with greater than or equal to 10 pack-year smoking history, with and without airflow obstruction, were enrolled between January 2008 and June 2011. Participants were non-Hispanic white or African American. Participants underwent volumetric inspiratory and expiratory CT using standardized protocol; images were transferred to a central lab for protocol verification and quality control (QC)^12^. Exclusion criteria included a history of other lung disease (except asthma), prior surgical excision involving a lung lobe or greater, present cancer, metal in the chest, or history of chest radiation therapy. Participants were excluded from the present study due to inadequate CT for computing tPRM, such as missing an inspiration/expiration scan, or failing QC implemented specifically for the present study. Our QC protocol is described in **Appendix 1**. Data for participants evaluated here have been utilized in numerous previous studies and a list of COPDGene publications can be found at http://www.copdgene.org/publications. Our study is the first to report tPRM analysis across the whole Phase 1 cohort and predict spirometric decline over 5 years in the Phase 2 subset of COPDGene participants.

Spirometry was performed in the COPDGene study before and after the administration of a bronchodilator, specifically 180 mcg of albuterol (Easy-One spirometer; NDD, Andover, MA). Post-bronchodilator values were used in our analyses. COPD was defined by a post-bronchodilator FEV_1_/FVC of less than 0.7 at the baseline visit, as specified in the Global Initiative for Chronic Obstructive Lung Disease (GOLD) guidelines^13^. GOLD grades 1-4 were used to define disease severity. GOLD 0 classification, i.e., “at-risk,” was defined by a post-bronchodilator FEV_1_/FVC ≥ 0.7 at the baseline visit, alongside FEV_1_% predicted ≥ 80%. Participants with FEV_1_/FVC ≥ 0.7 with FEV_1_% predicted < 80% were classified as having preserved ratio impaired spirometry (PRISm)^14^. Demographic and spirometric measures used in this study included age, sex, race, smoking history, scanner manufacturer, body mass index (BMI), FEV_1_% predicted, FEV_1_/FVC and forced mid-expiratory flow (FEF_25-75_).

### Computed Tomography and Topological PRM Analysis

All computed tomography (CT) data were obtained from multiple sites associated with the COPDGene project at Phase 1. Whole-lung volumetric multidetector CT acquisition was performed at full inspiration and normal expiration at functional residual capacity using a standardized previously published protocol^12^. Data reconstructed with the standard reconstruction kernel was used for quantitative analysis. All CT data were presented in Hounsfield units (HU), where stability of CT measurement for each scanner was monitored monthly using a custom COPDGene phantom^12^. For reference, air and water attenuation values are −1,000 and 0 HU, respectively.

PRM were determined from paired CT scans using Lung Density Analysis (LDA) software (Imbio, LLC, Minneapolis, MN). LDA segmented the lungs from the thoracic cavity with airways removed. Inspiratory CT scans were spatially aligned to the expiratory geometric frame using deformable image registration. Lung voxels were classified using pre-determined HU thresholds as: normal (PRM^Norm^, -950 < inspiration HU ≤ -810, and expiration HU ≥ -856), functional small airways disease (PRM^fSAD^, -950 < inspiration HU ≤ -810, expiration HU < -856), emphysema (PRM^Emph^, inspiration HU < -950, expiration HU < -856), or parenchymal disease (PRM^PD^, inspiration HU > -810)^15^. Only voxels between -1,000 HU and -250 HU at both inspiration and expiration were used for PRM classification. Each PRM classification was quantified as the percent volume, which is defined as the sum of a PRM classification normalized to the total lung volume at expiration multiplied by 100.

Topological analysis of PRM was performed using methods previously described^10^. tPRM metrics were defined through application of Minkowski measures on 3D binary voxel distributions: volume density (V) and Euler-Poincaré Characteristic (χ)^16^. Maps of V and χ were computed for each PRM class (Norm, fSAD, Emph, and PD) using a 3D moving window of size 21 x 21 x 21 voxels evaluated on a grid of every 5th voxel. V was normalized by the Minkowski estimate of the mask within the same local window volume (rather than a direct calculation of the mask volume in the window as previously described) and χ by the masked window voxel count. Linear interpolation was applied to determine V and χ values for all segmented voxels.

To indicate the PRM class associated with a Minkowski measure, the class is presented as a superscript (e.g., V^fSAD^ is the volume density of PRM^fSAD^). tPRM analysis was performed using open-source and in-house software developed in MATLAB R2019a (MATLAB, The MathWorks Inc., Natick, MA). A detailed overview and diagram, of computing tPRM from raw imaging data, was made by Hoff et al.^10^. Because the focus of this study is the relationship between normal parenchyma and SAD, and its association with emphysema, all analyses were performed using V and χ for PRM classifications Norm and fSAD. For completeness, V and χ for PRM classifications Emph and PD are provided.

### Phase 1 Data and Statistical Analysis

Data in this study are presented as mean and standard deviation unless stated otherwise. Correlation between V and χ for PRM^Norm^ and PRM^fSAD^ were calculated using Spearman rank-order correlation coefficients (*ρ*). The total Phase 1 cohort was separated into two subsets based on spirometry confirmed COPD: non-COPD (FEV_1_/FVC ≥ 0.7) and COPD (FEV_1_/FVC < 0.7). Cross-sectional multivariable regression analysis was performed on both subsets using a stepwise approach with V and χ for PRM classifications Norm and fSAD as independent variables and selected pulmonary function testing and clinical features as outcome variables, controlling for age, gender, race, BMI, smoking (pack years) and CT vendor. These control variables were included as compulsory independent variables in all regression models. Statistical work was conducted using IBM SPSS Statistics v27 (SPSS Software Products). In all tests, significance was defined by p < 0.05.

### Predict Spirometric Decline

We evaluated baseline V and χ for PRM classifications Norm and fSAD as predictors of FEV_1_ decline over 5 years using a machine learning (ML) model. A total of 4483 cases from the Phase 2 cohort of the COPDGene longitudinal trial, a subset of Phase 1, had FEV_1_ measurements at baseline and 5-year follow up. Our ML model is a sparse dictionary learning algorithm^17–20^ that classifies image patch features as “normal” or “abnormal”. In our method, we used the tPRM maps V^Norm^, V^fSAD^, χ^Norm^, and χ^fSAD^ of each case as inputs for training and testing the algorithm. For training our ML model, individual cases were stratified based on the change in FEV_1_ over 5 years [= (FEV_1_ at yr 5 – FEV_1_ at yr 0)/5 years] as fast (ΔFEV_1_/yr ≤ - 60ml/yr; N=1516) and slow progressors (ΔFEV_1_/yr > -60ml/yr; N=2967). We used 35% of the data for training and 65% for testing the model. Training was performed on a randomly selected subset of 1569 cases, with N = 531 fast progressors and N = 1038 slow progressors. The remaining 2914 cases, consisting of N = 985 fast progressors and N = 1929 slow progressors, were used for testing the algorithm. In brief, our ML model is designed to associate unique features from the input image patches with fast and slow progressors. This is achieved by randomly selecting image patches from within the lung and extracting the information from the inputs (tPRM maps V^Norm^, V^fSAD^, χ^Norm^, and χ^fSAD^ given as input to the ML algorithm) at these image patch locations and comparing their underlying patch features with the compiled class dictionaries of features, which are determined during training. It is important to note that no previous knowledge about the case and lung tissue features, such as emphysema, are provided for the algorithm to delineate “normal” from “abnormal” lung tissue. Details on model design and methods for training and testing are provided in **Figure 1** and **Appendix 2**. To determine the contribution of each feature to the model selection, we used the minimum redundancy maximum relevance feature selection algorithm^21^ to rank the tPRM inputs used in the dictionary learning algorithm. The algorithm quantifies the redundancy and relevance using mutual information of variables.^22, 23^ We also investigated the selection bias for each input in the ML model and obtained the prediction accuracy for 10 different choices of training image patches, considering each input separately in the model. The prediction accuracy for each training run is fit to a Gaussian probability density function^24^. All processing and analyses were performed using in-house algorithms developed in MATLAB version 2020a (MathWorks, Natick, MA). To determine the contribution of our ML model to account for spatial features in predicting FEV_1_ decline, we determined if whole lung mean values of V^Norm^, V^fSAD^, χ^Norm^, and χ^fSAD^ were as predictive of FEV_1_ decline using a logistic regression classifier.

**Figure 1:**
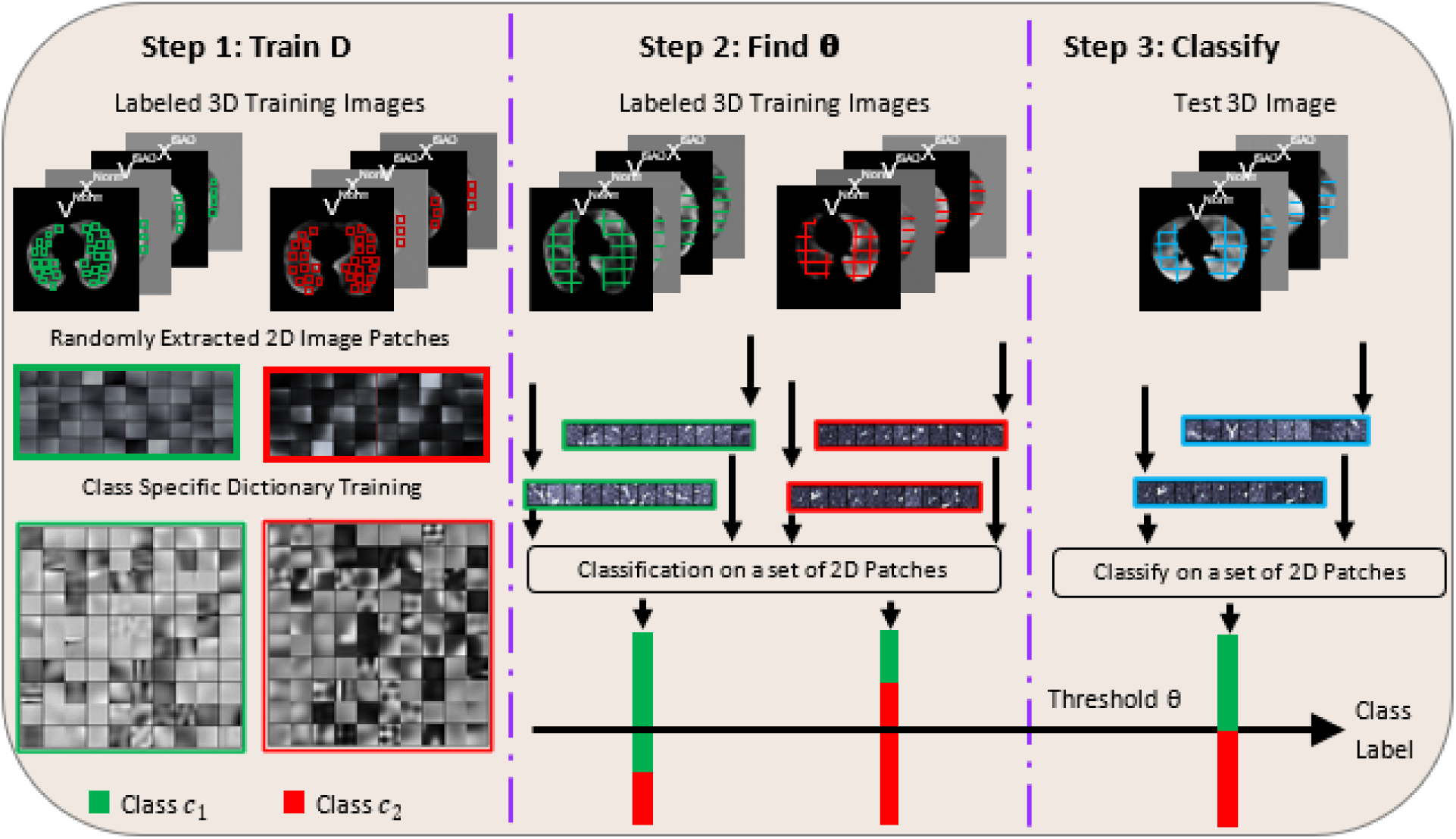
Flowchart describing the various steps involved in the proposed dictionary learning algorithm. Step 1: 2D image patches are extracted from each of the prior maps (V^Norm^, V^fSAD^, χ^Norm^, and χ^fSAD^) in the labeled training data and a class specific dictionary is trained for the entire class. Step 2: all 2D patches from each 3D prior image map are classified and a threshold value is selected to classify the entire case as belonging to one of the classes. Step 3: The learned class specific dictionaries and the threshold for the entire case are used to classify the test images.

### Case Study: Spatial Analysis

To better understand the relationship between PRM^fSAD^ and PRM^Emph^, we evaluated the spatial dependance of V and χ for these PRM classifications from a single subject. The case is a female subject, 45-50 years of age, diagnosed with GOLD 4 COPD. On a single axial slice, profiles of V and χ for PRM^fSAD^ and PRM^Emph^ were generated by selecting points from high emphysema (V^Emph^ > 0.6) and low emphysema (V^Emph^ < 0.2). A line plot (**Figure 2**) was produced for V and χ vs distance along each point of the profile. The distance, in units of centimeter, along the image profile was determined using the voxel dimensions of the CT scan. All processing and analyses were performed using in-house algorithms developed in MATLAB version 2020a (MathWorks, Natick, MA).

**Figure 2:**
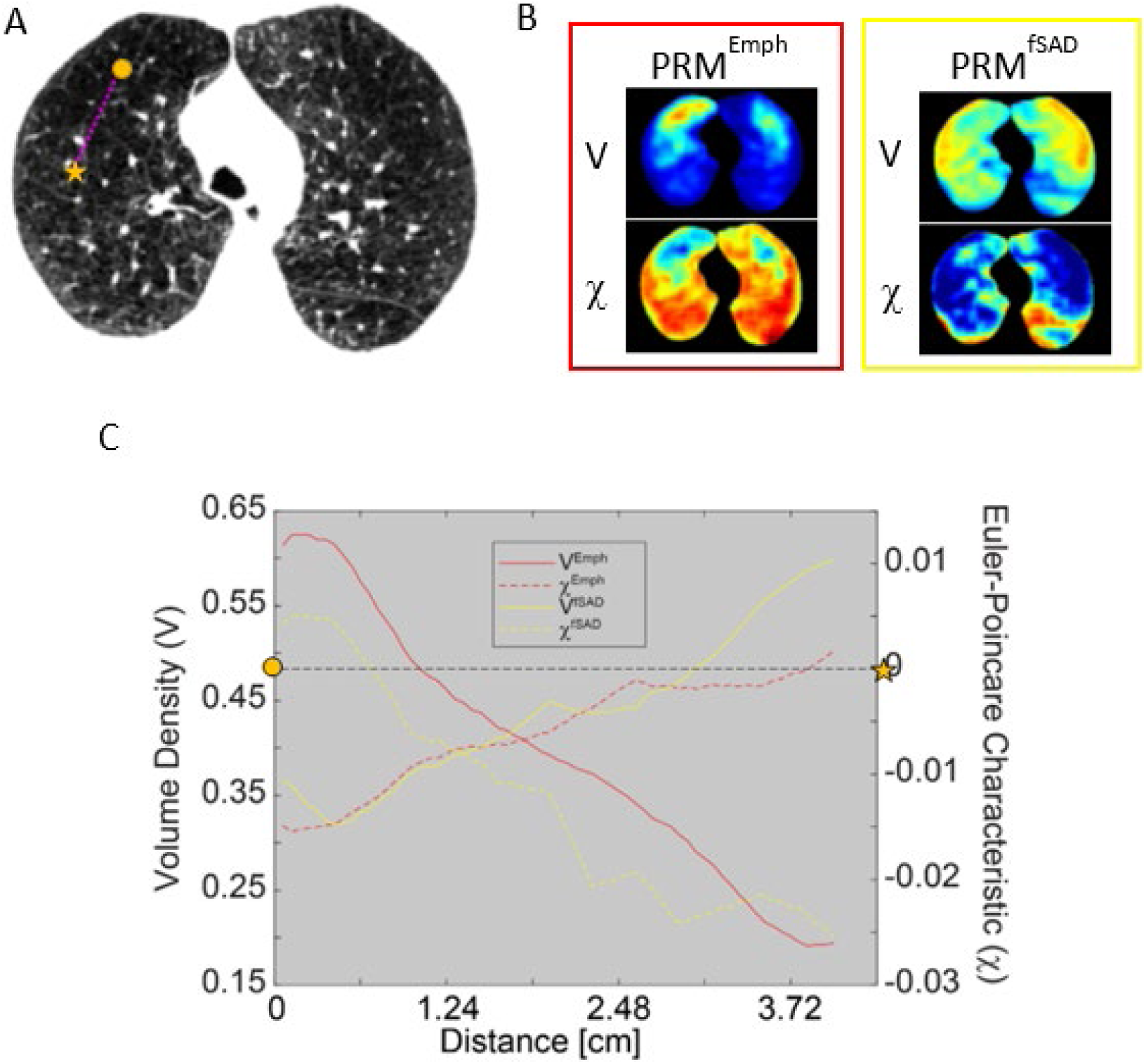
Case study demonstrating the spatial relationship between the topologies of PRM^fSAD^ and PRM^Emph^. The case is a female subject, 45-50 years of age, diagnosed with GOLD 4 COPD. Single axial slice from (A) spatially aligned CT scan acquired at full inflation with corresponding (B) slices from V and χ of PRM^fSAD^ and PRM^Emph^. (C) Topology values were plotted along the dashed line on the CT slice, starting from circle to star. Lines on plot were color coded to match PRM classification (red signifies PRM^Emph^ and yellow signifies PRM^fSAD^). Solid and dashed lines indicate V (left y-axis) and χ (right y-axis).

## Results

### Population Characteristics

The original COPDGene Phase 1 cohort consisted of 10,300 individuals. We excluded 1,344 participants for: inadequate CT data, such as missing an expiration or inspiration scan, to conduct tPRM analysis (n = 1,125); missing clinical data (n = 16); or failing to pass our CT-based QC testing (n = 203). Further details of CT QC are provided in **Appendix 1**. The resulting complete subset used for analyses thus consisted of 8,956 participants. Baseline demographics and lung function for all Phase 1 participants, grouped based on FEV_1_% predicted and FEV_1_/FVC—that is, by GOLD grade or PRISm as described in the Materials and Methods—are reported in **Table 1**. Due to the COPDGene recruitment strategy, the proportion of GOLD 0 (FEV_1_/FVC ≥ 0.7, FEV_1_% predicted ≥ 80%) participants^12^ account for almost half of the study population (43%; 3,867 of 8,956 participants). Increasing percent volume of PRM-derived fSAD (PRM^fSAD^) and PRM-derived emphysema (PRM^Emph^), with decreasing PRM^Norm^, was observed with higher GOLD grades. This is consistent with previously published work. PRM-derived parenchymal disease (PRM^PD^) was found to be elevated in PRISm and GOLD 0 participants (35.8 ± 16.4% and 26.3 ± 12.8% of the total lung volume, respectively) as compared to the COPD subset.

**Table 1:**
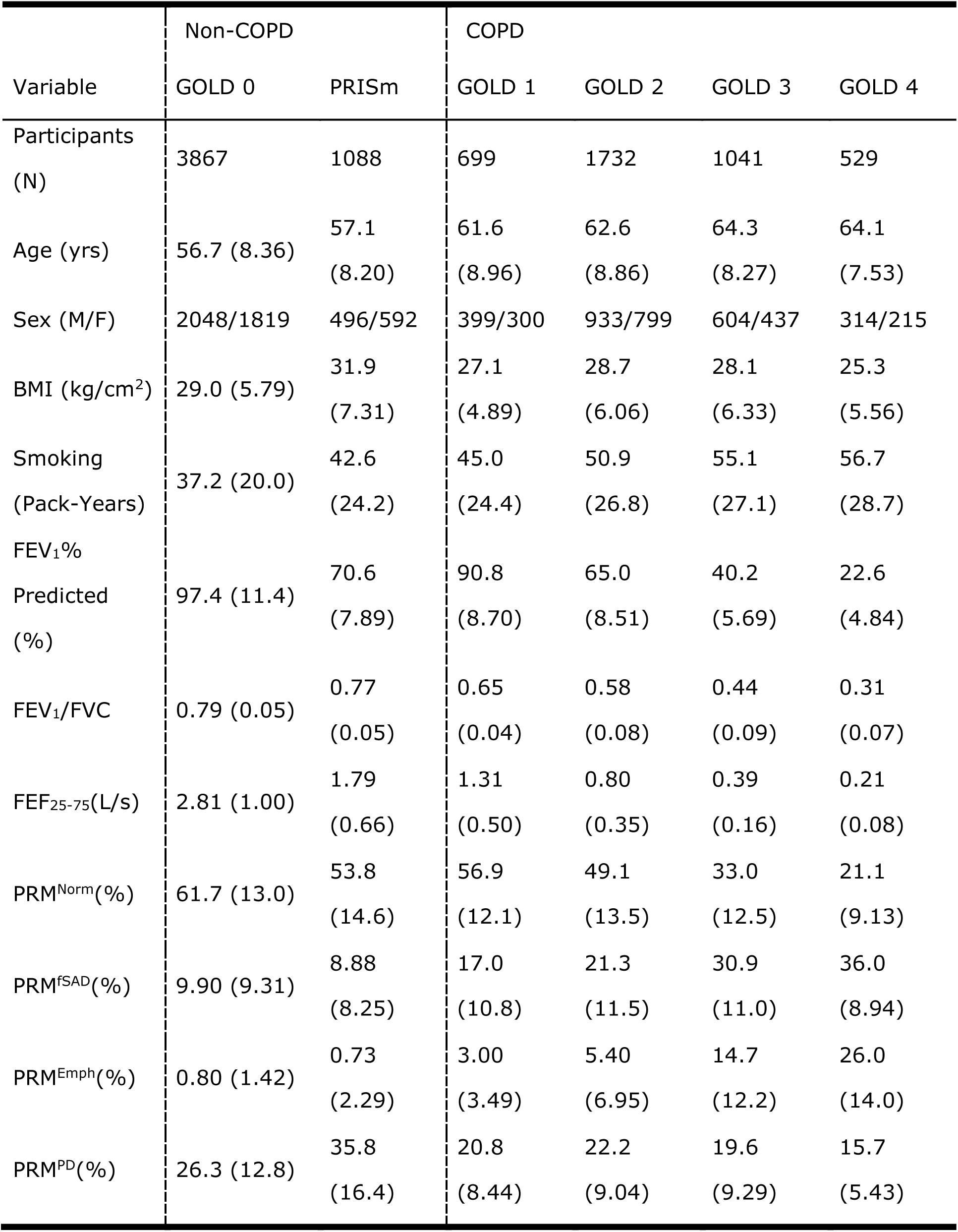
Clinical Characterization of the Study Population Notes. : Participant characteristics of the entire study population separated in subsets of those with (FEV_1_/FVC < 0.7) and without (FEV_1_/FVC ≥ 0.7) COPD. Values are displayed as mean (standard deviation). GOLD, Global Initiative for Chronic Obstructive Lung Disease; PRISm, preserved ratio impaired spirometry; GOLD 0, at-risk smokers with normal spirometry; BMI, body mass index; FEV_1_, forced expiratory volume in one second; FVC, forced vital capacity; FEF_25-75_, forced mid-expiratory flow; PRM, parametric response map; Norm, Normal; fSAD, functional small airways disease; Emph, emphysema; PD, parenchymal disease.

| Variable | Non-COPD |  | COPD |  |  |  |
| --- | --- | --- | --- | --- | --- | --- |
|  | GOLD 0 | PRISm | GOLD 1 | GOLD 2 | GOLD 3 | GOLD 4 |
| Participants (N) | 3867 | 1088 | 699 | 1732 | 1041 | 529 |
| Age (yrs) | 56.7 (8.36) | 57.1 (8.20) | 61.6 (8.96) | 62.6 (8.86) | 64.3 (8.27) | 64.1 (7.53) |
| Sex (M/F) | 2048/1819 | 496/592 | 399/300 | 933/799 | 604/437 | 314/215 |
| BMI (kg/cm<sup>2</sup>) | 29.0 (5.79) | 31.9 (7.31) | 27.1 (4.89) | 28.7 (6.06) | 28.1 (6.33) | 25.3 (5.56) |
| Smoking (Pack-Years) | 37.2 (20.0) | 42.6 (24.2) | 45.0 (24.4) | 50.9 (26.8) | 55.1 (27.1) | 56.7 (28.7) |
| FEV<sub>1</sub>% Predicted (%) | 97.4 (11.4) | 70.6 (7.89) | 90.8 (8.70) | 65.0 (8.51) | 40.2 (5.69) | 22.6 (4.84) |
| FEV<sub>1</sub>/FVC | 0.79 (0.05) | 0.77 (0.05) | 0.65 (0.04) | 0.58 (0.08) | 0.44 (0.09) | 0.31 (0.07) |
| FEF<sub>25-75</sub>(L/s) | 2.81 (1.00) | 1.79 (0.66) | 1.31 (0.50) | 0.80 (0.35) | 0.39 (0.16) | 0.21 (0.08) |
| PRM<sup>Norm</sup>(%) | 61.7 (13.0) | 53.8 (14.6) | 56.9 (12.1) | 49.1 (13.5) | 33.0 (12.5) | 21.1 (9.13) |
| PRM<sup>fSAD</sup>(%) | 9.90 (9.31) | 8.88 (8.25) | 17.0 (10.8) | 21.3 (11.5) | 30.9 (11.0) | 36.0 (8.94) |
| PRM<sup>Emph</sup>(%) | 0.80 (1.42) | 0.73 (2.29) | 3.00 (3.49) | 5.40 (6.95) | 14.7 (12.2) | 26.0 (14.0) |
| PRM<sup>PD</sup>(%) | 26.3 (12.8) | 35.8 (16.4) | 20.8 (8.44) | 22.2 (9.04) | 19.6 (9.29) | 15.7 (5.43) |

### Topological Readouts of PRM

Presented in **Figure 3** is a case with elevated fSAD (PRM^fSAD^ = 40%). Representative coronal slices of the expiration CT scan and PRM^fSAD^, overlaid on CT scan, are provided. To illustrate the dependence of V and χ on the arrangement of PRM^fSAD^, we have included V^fSAD^ and χ^fSAD^ maps indicating regions with low (cyan box) and high (magenta box) V^fSAD^. As expected, V^fSAD^ (**Figure 3C**) is dependent on the amount of fSAD (yellow voxels in **Figure 3B**). Averaged over the lungs, V^fSAD^ is proportional to the percent volume of PRM^fSAD^ by a factor of 100. However, χ^fSAD^ > 0 (cyan box in **Figure 3D**) corresponds to the formation of fSAD pockets (cyan box **Figure 3B**), whereas χ^fSAD^ < 0 (cyan box in **Figure 3D**) is the consolidation of these pockets into a mesh with holes (magenta box in **Figure 3B**).

**Figure 3:**
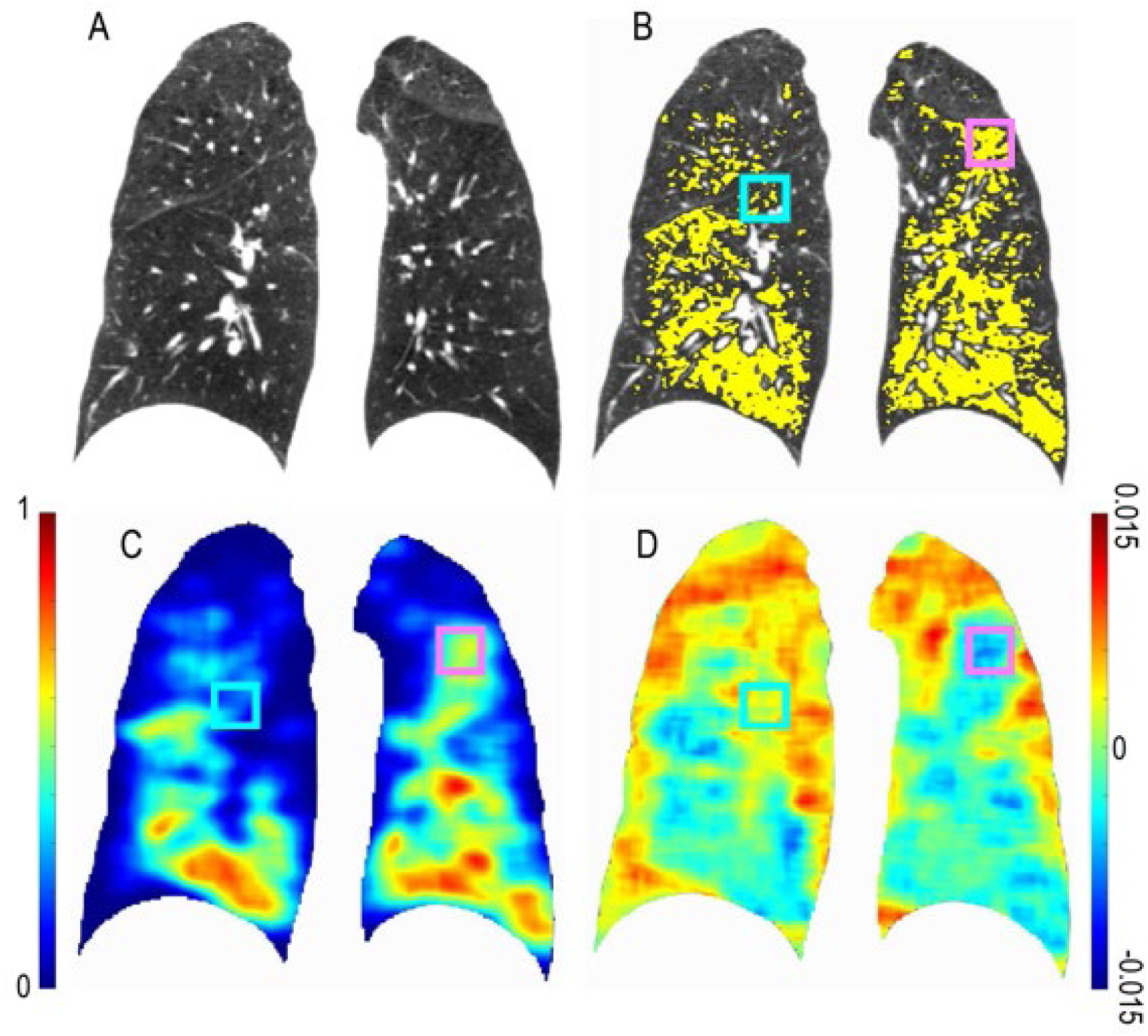
Illustration of Volume Density (V) and Euler-Poincaré Characteristic (χ) for PRM^fSAD^. Presented are representative coronal slices for the (A) expiratory CT scan with associated (B) PRM^fSAD^ overlay (yellow). Included are the (C) volume density and (D) Euler-Poincaré Characteristic of PRM^fSAD^. Magenta box indicates a lung region with elevated V^fSAD^ and negative χ^fSAD^. Cyan box indicates a lung region with minimal V^fSAD^ and positive χ^fSAD^. The subject is a GOLD 3 female, 50-55 years of age, with FEV_1_% predicted of 32% and percent volume of PRM^fSAD^ of 40%.

The volume density of PRM^Norm^ and PRM^fSAD^ demonstrated an inverse relationship with increasing COPD severity (**Figure 4A**), consistent with previous work. A similar inverse relationship was observed for χ of both normal lung and fSAD (χ^Norm^ and χ^fSAD^). Values of χ^Norm^ and χ^fSAD^ were found to flip about zero (e.g., χ^fSAD^ changes from positive to negative values) from GOLD 2 to GOLD 4 (**Figure 4B**). As provided in **Table 2**, χ^Norm^ and χ^fSAD^ had means (standard deviations) of -0.0084 (0.0071) and 0.0047 (0.0074), respectively, for cases diagnosed as GOLD 2. For those with severe COPD, i.e., GOLD 4, χ^Norm^ and χ^fSAD^ were 0.0039 (0.0055) and -0.0036 (0.0048), respectively. Mean values of χ^Emph^ and χ^PD^ were found to be positive and similar across GOLD (**Table 2**).

**Figure 4:**
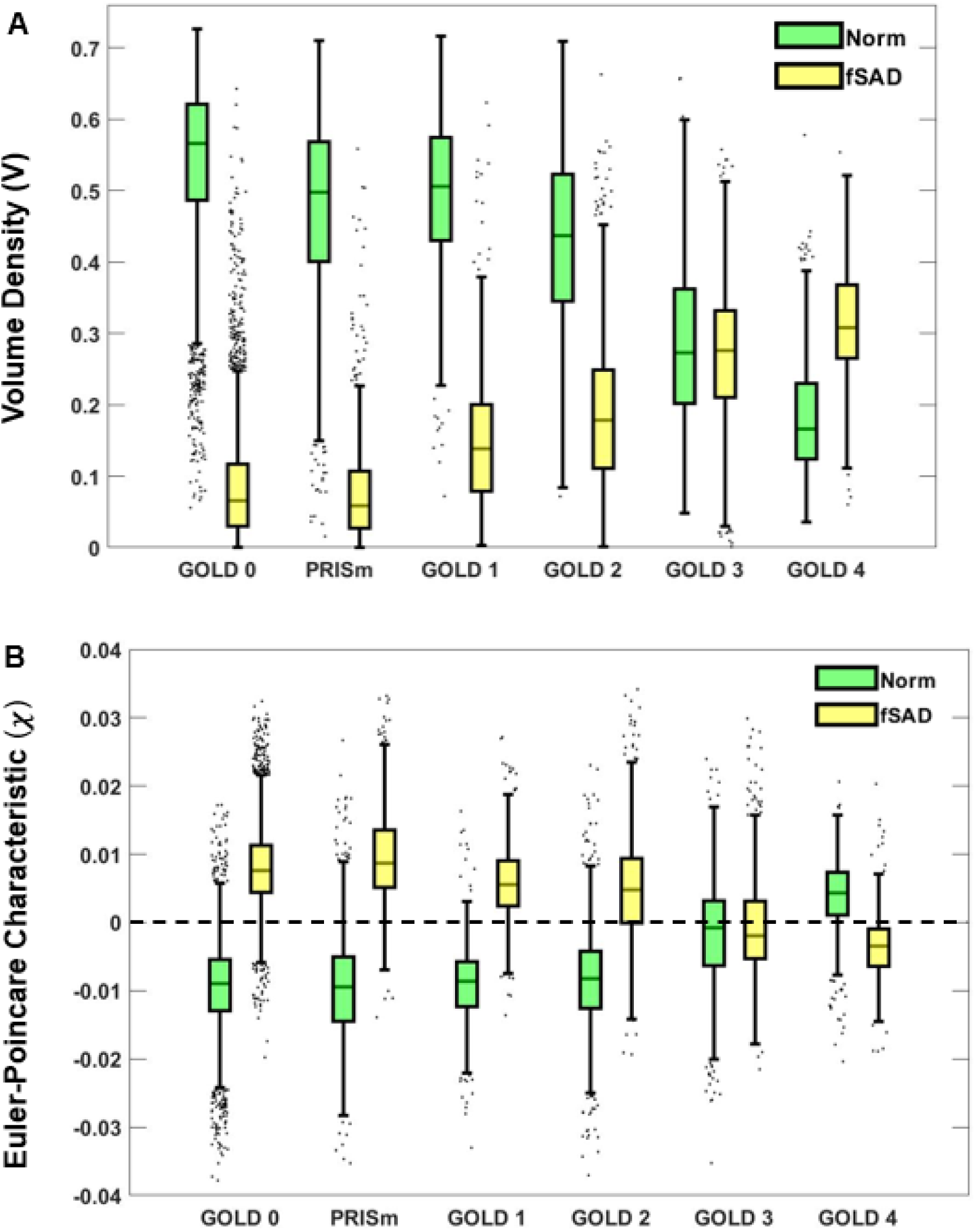
Boxplots for Volume Density (V) and Euler-Poincaré Characteristic (χ) of PRM^Norm^ and PRM^fSAD^ across all GOLD stages, “at-risk” (GOLD 0) and PRISm. Plots of (A) V and (B) χ are provided for PRM^Norm^ (green) and PRM^fSAD^ (yellow). Box plots were computed following standard protocol for box and whiskers. Box lines determined by lower quartile (Q1), middle quartile / median (Q2), and upper quartile (Q3). Whiskers are drawn out to Q1 - 1.5 x IQR and Q3 + 1.5 x IQR for lower and upper limits respectively. IQR = Q3-Q1. Outliers are defined as points beyond the given upper and lower limits and illustrated as black points with a random bounded horizontal perturbation beyond box whiskers.

**Table 2:** Topology Readouts for all PRM Classifications Notes: Data are presented as the mean (standard deviation). The entire study population was separated into subsets of those with (FEV_1_/FVC < 0.7) and without (FEV_1_/FVC ≥ 0.7) COPD. GOLD, Global Initiative for Chronic Obstructive Lung Disease; PRISm, preserved ratio impaired spirometry; GOLD 0, at-risk smokers with normal spirometry; PRM, Parametric Response Map; Norm, normal lung parenchyma; fSAD, functional small airways disease; Emph, emphysema; and PD, parenchymal disease.

| Topology | PRM | Non-COPD |  | COPD |  |  |  |
| --- | --- | --- | --- | --- | --- | --- | --- |
|  |  | GOLD 0 | PRISm | GOLD 1 | GOLD 2 | GOLD 3 | GOLD 4 |
| Volume Density [V] | Norm | 0.62 (0.13) | 0.54 (0.15) | 0.57 (0.12) | 0.5 (0.14) | 0.33 (0.13) | 0.21 (0.09) |
|  | fSAD | 0.1 (0.09) | 0.09 (0.08) | 0.17 (0.11) | 0.22 (0.12) | 0.31 (0.11) | 0.36 (0.09) |
|  | Emph | 0.01 (0.01) | 0.01 (0.02) | 0.03 (0.04) | 0.05 (0.07) | 0.15 (0.12) | 0.26 (0.14) |
|  | PD | 0.25 (0.13) | 0.35 (0.16) | 0.2 (0.08) | 0.21 (0.09) | 0.19 (0.09) | 0.15 (0.05) |
| Euler-Poncairé<br>Characteristic [$\chi$] | Norm | -0.0095 (0.0063) | -0.0095 (0.008) | -0.0091 (0.0055) | -0.0084 (0.0071) | -0.0016 (0.0078) | 0.0039 (0.0055) |
|  | fSAD | 0.0081 (0.0059) | 0.0096 (0.0065) | 0.0058 (0.0057) | 0.0047 (0.0074) | -0.0009 (0.0071) | -0.0036 (0.0048) |
|  | Emph | 0.0024 (0.003) | 0.0018 (0.0025) | 0.0046 (0.0039) | 0.005 (0.0039) | 0.0052 (0.0039) | 0.0033 (0.0032) |
|  | PD | 0.0044 (0.0033) | 0.0028 (0.0046) | 0.0043 (0.0018) | 0.004 (0.0022) | 0.0031 (0.0017) | 0.0024 (0.0009) |

**Notes:** Data are presented as the mean (standard deviation). The entire study population was separated into subsets of those with ( $FEV_1/FVC < 0.7$ ) and without ( $FEV_1/FVC \geq 0.7$ ) COPD. GOLD, Global Initiative for Chronic Obstructive Lung Disease; PRISm, preserved ratio impaired spirometry; GOLD 0, at-risk smokers with normal spirometry; PRM, Parametric Response Map; Norm, normal lung parenchyma; fSAD, functional small airways disease; Emph, emphysema; and PD, parenchymal disease.

We further evaluated the relationship of PRM^Norm^ and PRM^fSAD^ with respect to V (**Figure 5A**) and χ (**Figure 5B**). Both V and χ demonstrated strong correlations between Norm and fSAD (ρ = -0.666, p < 0.001 and ρ = -0.745, p < 0.001, respectively) over the Phase 1 cohort. Here the GOLD stages are coded by color and the relative amount of emphysema, quantified by V^Emph^, by size of the marker. As observed in **Figure 5A**, V^Norm^ versus V^fSAD^ had more scatter in the data compared to χ^Norm^ versus χ^fSAD^ (**Figure 5B**). As expected, GOLD 4 cases with elevated emphysema (V^Emph^) demonstrated a drop in V^Norm^ and V^fSAD^ values. In contrast, χ^Norm^ consisted of primarily positive values, whereas positive and negative values were observed for χ^fSAD^ (**Figure 5B**). Although V^fSAD^ was found to be strongly correlated to V^Emph^ (r = 0.845, p < 0.001), only a weak correlation was observed between χ^fSAD^ and χ^Emph^ (ρ = -0.155, p < 0.001).

**Figure 5:**
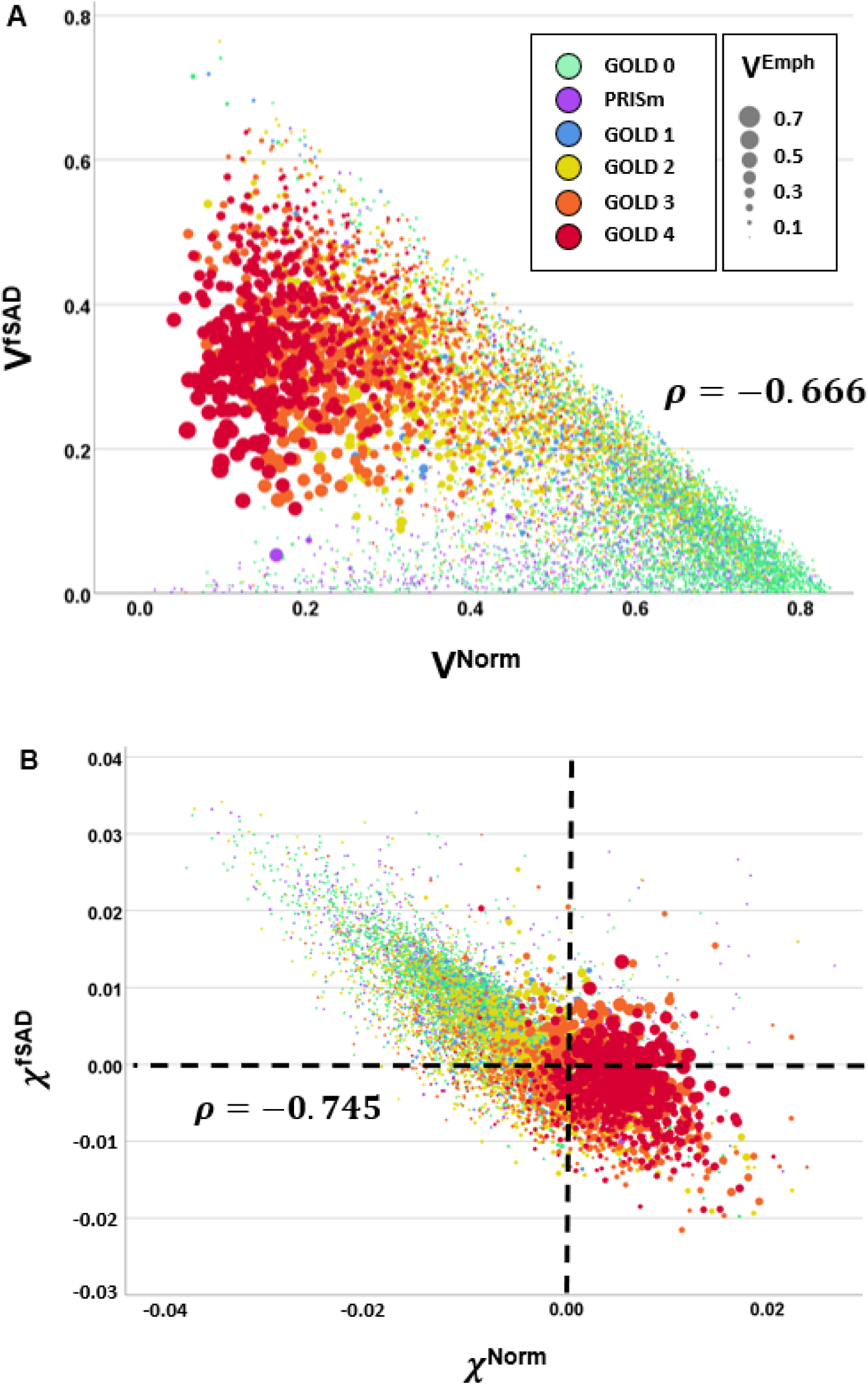
Scatter plots of all study sample participants for (A) V^Norm^ versus V^fSAD^ and (B) χ^Norm^ versus χ^fSAD^. Individual points are color coded based on COPD classifications. The size of the points indicates the amount of emphysema as measured by the volume density of PRM^Emph^ (V^Emph^).

### Multivariable Regression Analysis

Presented in **Table 3** are results from multivariable regression analyses that demonstrate the contribution of V and χ to PRM^Norm^ and PRM^fSAD^ when modeling spirometric measures and the volume density of emphysema, controlling for age, sex, race, BMI, pack-years, and CT vendor. Among those with spirometrically confirmed COPD, V^Norm^ was found to be significantly associated with multiple clinical measures including FEV_1_% predicted, FEV_1_/FVC, FEF_25-75_ and V^Emph^ (see **Table 3**). V^fSAD^ and χ^fSAD^ were found to independently and significantly contribute to FEV_1_% predicted (β = 0.065, p=0.004 and β = 0.106, p<0.001). Only the Norm topological measures were found to contribute to FEV_1_/FVC (β = 0.668, p<0.001 for V^Norm^ and β = - 0.120, p<0.001 for χ^Norm^), whereas V and χ for both Norm and fSAD were found to be significant parameters for FEF_25-75_. With respect to V^Emph^, extent of emphysema, V and χ for Norm and fSAD were highly significant but demonstrated similar trends irrespective of PRM classification. For completeness, the same analyses were performed on the non-COPD cohort (**Table 4**). As compared to the COPD cohort, statistical models generated from the non-COPD cohort demonstrated significant parameters but with weaker correlations (i.e., adjusted R^2^).

**Table 3:** Multivariable Regression for COPD Subset Notes: Multivariable regression modelling using volume density (V) and Euler-Poincaré Characteristic (χ) for PRM-derived Normal and fSAD (introduced stepwise) to model pulmonary function testing measures in the COPD subset. Each column presents results for a different regression model. FEV_1_, forced expiratory volume in one second; FVC, forced vital capacity; FEF_25-75_, forced mid-expiratory flow; Emph, emphysema; SE, standard error of the estimate; BMI, body mass index; Norm, Normal; fSAD, functional small airways disease. Model performance is reported as adjusted R^2^ and standard error of the estimate. Feature association is reported as standardized beta coefficients (β); cells for stepwise variables removed from final model. All regression models were controlled for age, sex, race, BMI, pack years and CT vendor. P values ≥ 0.01, < 0.01 and ≥ 0.001, and < 0.001 are presented as values in parentheses, *, and **, respectively.

| Performance | FEV<sub>1</sub>% predicted | FEV<sub>1</sub>/FVC | FEF<sub>25-75</sub> (L) | V<sup>Emph</sup> |
| --- | --- | --- | --- | --- |
| Adjusted R<sup>2</sup> | 0.516 | 0.602 | 0.526 | 0.778 |
| SE | 15.8 | 0.084 | 0.331 | 0.057 |
| Age (yrs) | 0.085** | 0.021 (0.06) | -0.184** | 0.035** |
| Sex (M/F) |  | 0.018 (0.08) | -0.283** | -0.035** |
| BMI (kg/cm<sup>2</sup>) | -0.110** | 0.033* |  | -0.232** |
| Smoking (Pack Years) | -0.046** | -0.013 (0.22) | -0.051** | -0.015 (0.06) |
| CT vendor |  |  |  | 0.111** |
| Race |  | 0.113** | -0.033* |  |
| V<sup>Norm</sup> | 0.727** | 0.668** | 0.688** | -1.01** |
| V<sup>fSAD</sup> | 0.065* |  | 0.138** | -0.408** |
| χ<sup>Norm</sup> |  | -0.120** | 0.134** | 0.150** |
| χ<sup>fSAD</sup> | 0.106** |  | 0.175** | 0.118** |

**Table 4:** Multivariable Regression for non-COPD Subset Notes: Multivariable regression modelling using volume density (V) and Euler-Poincaré Characteristic (χ) for PRM-derived Normal and fSAD (introduced stepwise) to model pulmonary function test measures in the COPD subset. Each column presents results for a different regression model. FEV_1_pp, forced expiratory volume in one second percent predicted; FEV_1_, forced expiratory volume in one second; FVC, forced vital capacity; FEF_25-75_, forced mid-expiratory flow; Emph, emphysema; SE, standard error of the estimate; BMI, body mass index; Norm, normal; and fSAD, functional small airways disease. Model performance is reported as adjusted R^2^ and standard error of the estimate. Feature association is reported as standardized beta coefficients (β); cells for stepwise variables removed from final model. All regression models were controlled for age, sex, race, BMI, pack years and CT vendor. P values ≥ 0.01, < 0.01 and ≥ 0.001, and < 0.001 are presented as values in parentheses, *, and **, respectively.

| Performance | FEV<sub>1pp</sub> | FEV<sub>1</sub>/FVC | FEF<sub>25-75</sub> (L) | V<sup>Emph</sup> |
| --- | --- | --- | --- | --- |
| Adjusted R<sup>2</sup> | 0.143 | 0.126 | 0.265 | 0.263 |
| SE | 14.39 | 0.048 | 0.883 | 0.014 |
| Age (yrs) |  | -0.119** | -0.290** | 0.042* |
| Sex (M/F) | -0.027 (0.05) | 0.014 (0.298) | -0.381** | -0.062** |
| BMI (kg/cm<sup>2</sup>) | -0.075** | 0.104** |  | -0.130** |
| Smoking (Pack-Years) | -0.105** | -0.098** | -0.098** |  |
| CT vendor | -0.052** |  | -0.015 (0.252) | 0.185** |
| Race |  | 0.117** | -0.068** |  |
| V<sup>Norm</sup> | 0.325** | -0.044* | 0.109** | -0.157** |
| V<sup>fSAD</sup> | 0.134** | -0.218** | -0.075** | 0.373** |
| χ<sup>Norm</sup> |  |  |  | -0.038 (0.01) |
| χ<sup>fSAD</sup> | -0.105** | -0.075** | -0.058** |  |

### Prediction Model of Spirometric Decline

Representative axial slices of expiration CT scan, PRM, V^fSAD^, χ^fSAD^ and corresponding patch probability maps from a fast progressor (with ΔFEV_1_/yr of -249 ml/yr) are provided in **Figure 6**. Our ML model correctly classified this subject as a fast progressor. This case is a male, 60- 65 years of age, diagnosed at baseline with GOLD 2 COPD. Using V and χ from PRM^fSAD^ and PRM^Norm^ as inputs, the ML model was able to determine regions of emphysema, discernible from existing fSAD, observed in the right upper lung as “abnormal” (blue patches in the probability maps). In contrast, the dorsal lung regions were classified as “normal” (red patches in the probability maps) due to the absence of fSAD and emphysema. For completeness we have provided in **Figure 7** representative axial slices of expiration CT scan, PRM, V^fSAD^, χ^fSAD^ and corresponding patch probability maps from a slow progressor (with ΔFEV_1_/yr of 101 ml/yr).

**Figure 6:**
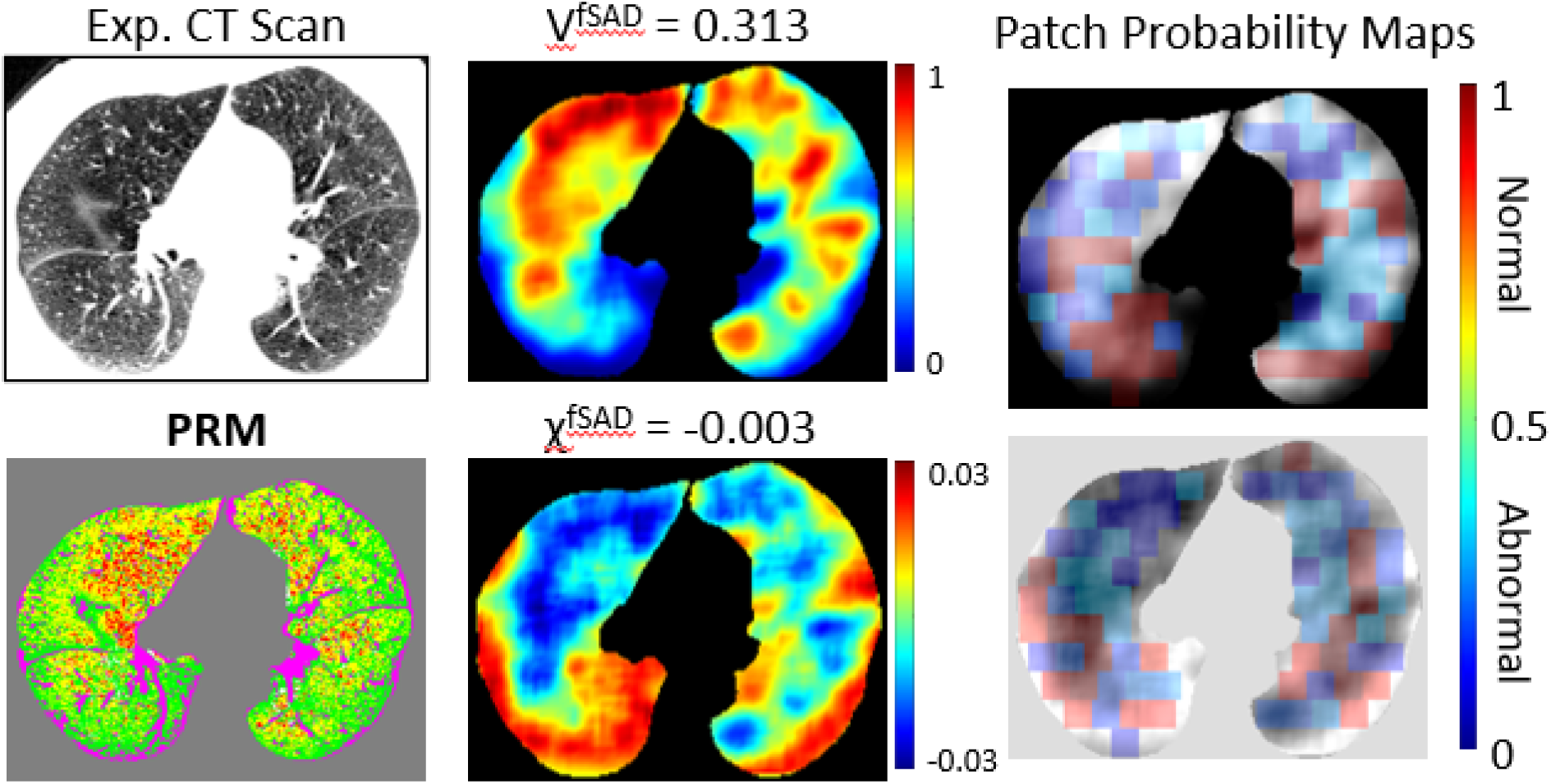
The dictionary learning results on a 60- to 65-year-old male diagnosed at baseline with GOLD 2 COPD and a fast progressor with ΔFEV_1_/yr of -249 ml/yr. Representative axial slice of an expiratory CT scan acquired at baseline, its associated PRM map, the tPRM maps V^fSAD^ and χ^fSAD^ of PRM^fSAD^, and their image patch probability maps from the dictionary learning model.

**Figure 7:**
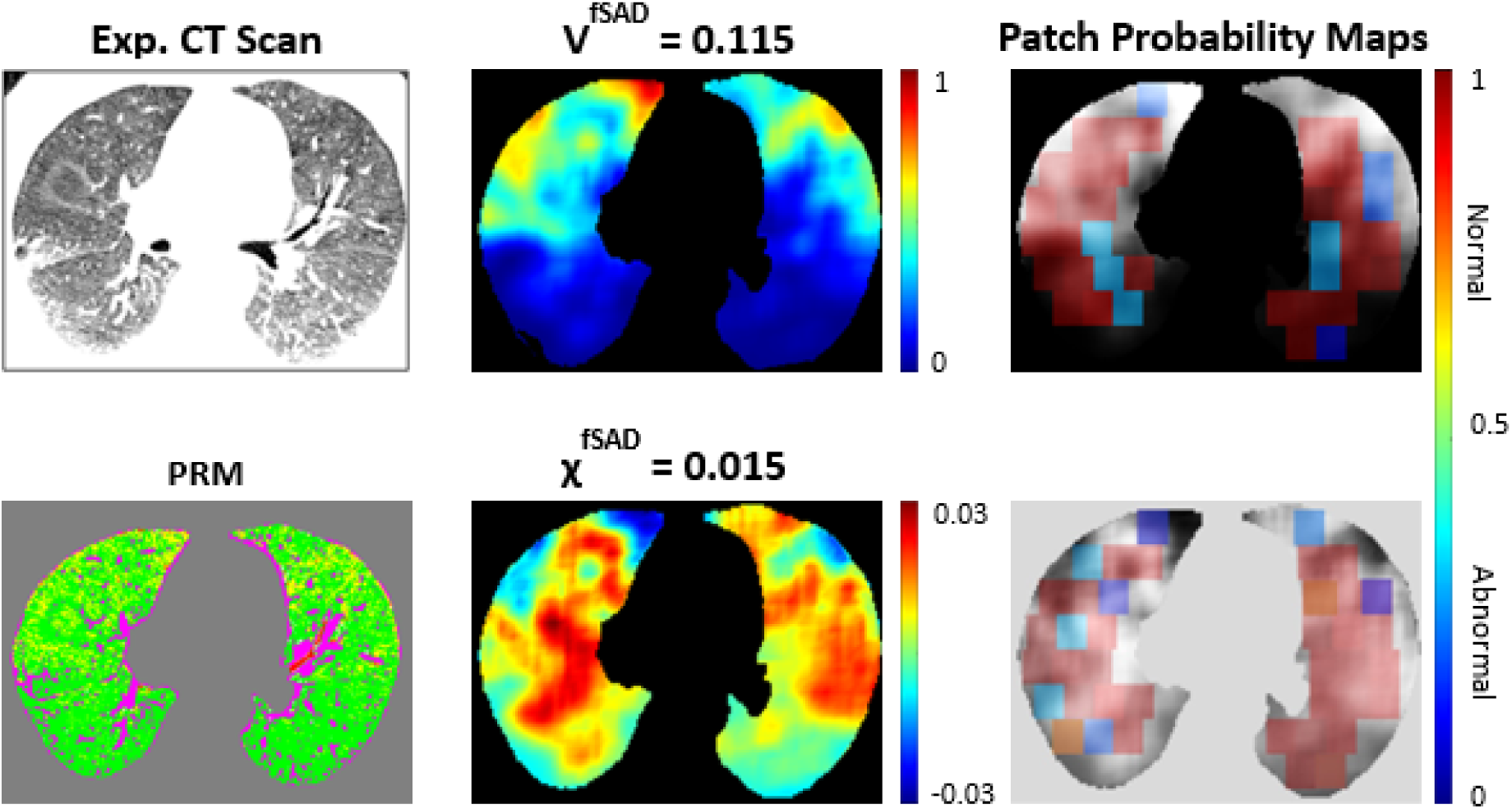
The dictionary learning results for a 70- to 75-year-old male diagnosed at baseline with GOLD 1 COPD and declared a slow progressor with ΔFEV_1_/yr of 101 ml/yr. This case was correctly identified by our ML algorithm as a slow progressor. Representative axial slice of an expiratory CT scan, its associated PRM map, the tPRM maps V^fSAD^ and χ^fSAD^ of PRM^fSAD^, and their image patch probability maps from the dictionary learning model.

As seen in **Figure 8A** and **B**, our ML model had an overall classification accuracy of 70.6% and Area Under the Curve (AUC) of 0.69 of the receiver operating characteristic (ROC) curve. We compared our ML model with a simple logistic regression model using whole lung mean values of V^Norm^, V^fSAD^, χ^Norm^, and χ^fSAD^. **Figure 8B** shows that the logistic regression model only achieved an AUC of 0.55. The contribution of each of the inputs to the model (V^Norm^, V^fSAD^, χ^Norm^, and χ^fSAD^) are shown in **Figure 8C** and **D**. V and χ of PRM^fSAD^ are dominant inputs, followed by V and χ of PRM^Norm^ (**Figure 8C**). Using a feature rank analysis performed on our test set, we observed that V and χ of PRM^fSAD^ are important to achieve higher prediction accuracy. In fact, χ^fSAD^ was found to have the smallest spread/variance (**Figure 8D**), indicating highly desirable robustness to the choice of training image patches and its usefulness as an input in the ML model. As reported in **Table 5**, “normal” patches, on average, consisted primarily of PRM^Norm^, elevated V^Norm^ (abundant) and negative χ^Norm^ (consolidated), with negligible PRM^fSAD^, low V^fSAD^ (depleted) and positive χ^fSAD^ (sparse pockets). In “abnormal” patches, similar values of V and χ for PRM^Norm^ and PRM^fSAD^ were observed (**Table 5**). Positive and negative values in χ^fSAD^ were found for “normal” and “abnormal” patches, respectively. This is consistent with the inverse relationship seen with increasing COPD severity shown in **Figure 4**.

**Figure 8:**
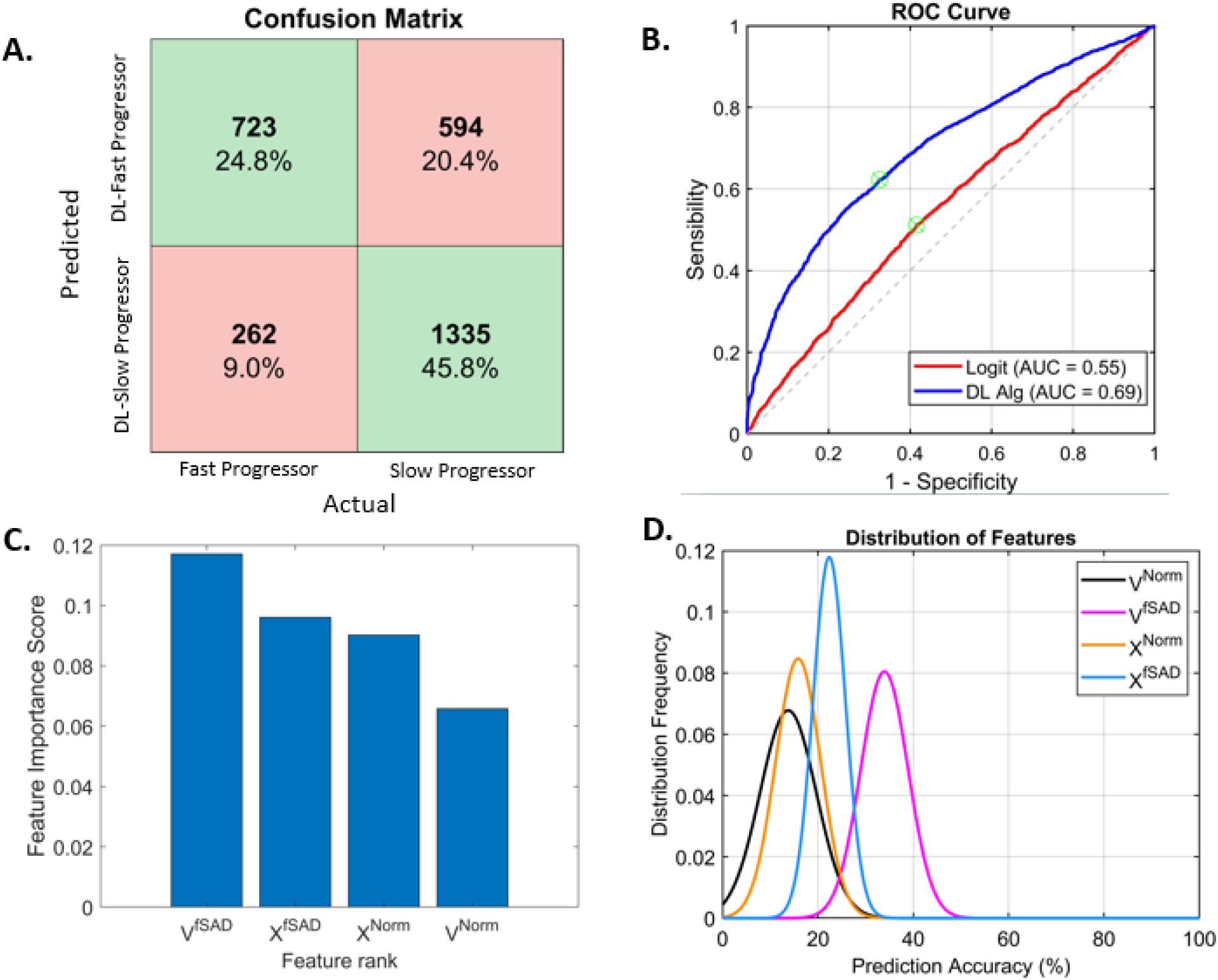
Results and relevance of the different features (tPRM metrics as inputs) used in the dictionary learning method. (A) Confusion Matrix showing the sensitivity and specificity of the ML model classifications for both the fast progressor (N = 985) and the slow progressor (N = 1929) classes in the test set. Green colored and red colored fields in the matrix represent agreement and disagreement, respectively, of the ML model with the actual decision. (B) Receiver Operating Characteristic (ROC) curve for our ML model and the logistic regression classifier with the corresponding Area Under the Curve (AUC) statistics. (C) Bar plot showing the feature importance score and feature ranking using the minimum redundancy maximum relevance method. (D) Plot showing the distribution of the features and their prediction accuracy over ten different training runs.

**Table 5:** Image Patch topological PRM metrics in the ML model Notes: Data are presented as the mean (standard deviation).

| tPRM Metrics | Normal | Abnormal |
| --- | --- | --- |
| $V^{\text{Norm}}$ | 0.5458 (0.1587) | 0.3798 (0.0897) |
| $V^{\text{fSAD}}$ | 0.1059 (0.0935) | 0.3299 (0.1008) |
| $\chi^{\text{Norm}}$ | -0.0065 (0.0084) | -0.0031 (0.0058) |
| $\chi^{\text{fSAD}}$ | 0.0041 (0.0061) | -0.0019 (0.0062) |

**Notes:** Data are presented as the mean (standard deviation).

### Dependence Between Topologies of PRM^fSAD^ and PRM^Emph^

As the topologies of PRM were determined as averages over the whole lungs, we provide a case study illustrating the relationship between V and χ of PRM^fSAD^ and PRM^Emph^ at the local level. Presented in **Figure 2** are the profiles of V and χ of PRM^fSAD^ and PRM^Emph^ from a region of the right lung with elevated and reduced V^Emph^ (orange circle and star, respectively, in **Figure 2A** and **C**). The case is a female subject, 45-50 years of age, diagnosed with GOLD 4 COPD. The subject was found to have on average high levels of V^fSAD^ (0.37) with relatively elevated V^Emph^ (0.1). Mean values for the whole lungs of χ were 0.008 and -0.009 for PRM^Emph^ and PRM^fSAD^, respectively. As seen in **Figure 2C**, V^fSAD^ increased while V^Emph^ decreased further from lung with the highest level of V^Emph^ (∼0.6 at orange circle in **Figure 2A** and **C**). At approximately 1.8 cm, volume densities between PRM^fSAD^ and PRM^Emph^ transitioned. In addition, χ^fSAD^ was found to increase with decreasing χ^Emph^ with transition occurring at ∼1.2cm.

## Discussion

The topological parametric response map is an extension of the well-established PRM method, a quantitative imaging marker of SAD^8^. In this study, we have demonstrated that inclusion of topological features, in this case the Euler-Poincaré Characteristic (χ), improved characterization and interpretation of fSAD in COPD as a complimentary readout of volume density (V), which is equivalent to traditional percent volume of PRM classifications^10^. This study also evaluated the role of PRM-defined normal parenchyma (PRM^Norm^) and fSAD (PRM^fSAD^) as lone indicators of COPD severity. We observed distinct patterns in topological metrics with respect to GOLD grades and identified a complete inversion in topology, characterized by Euler-Poincaré Characteristic χ, between normal lung and fSAD, in mid-to-late stages of COPD. We also found V and χ of PRM^Norm^ and PRM^fSAD^ to have statistically significant correlation with spirometric measures and emphysema and to be predictive of spirometric decline.

Our study builds on previous work by Hoff et al.^10^ on tPRM characterization in COPD. This study used a much smaller population (n = 88) to demonstrate the trends of all four topological features (volume density, surface area, mean curvature and Euler-Poincaré Characteristic) with increasing COPD severity^10^. Limited in statistical power, it instead focused on the surface area of fSAD. Access to a notably larger population (n = 8,956) in the current study allowed us to evaluate the volume density (V) and Euler-Poincaré Characteristic (χ) of PRM^Norm^ and PRM^fSAD^ and relate our findings to the field’s current understanding of COPD progression, i.e., normal parenchyma transitions to emphysema through SAD.

A key finding of our study is the ability to quantify parenchymal lung health, based not only on the extent but also on the arrangement of local lung abnormalities, i.e., fSAD. This is rooted in the concept that the lungs are healthy (i.e., PRM^Norm^) and COPD progresses through SAD (i.e., PRM^fSAD^), an intermediate between normal and emphysematous lung tissue, to emphysema. The nature of this transition suggests χ may be capturing a fundamental mechanism in the emergence of fSAD. Based on our observation, fSAD appears to develop as distinct pockets, which are represented as positive values in χ^fSAD^ within healthy lung tissue, as depicted in the cyan box in **Figure 3B**. With increasing COPD severity, fSAD pockets coalesce to a mesh, which is represented by negative values in χ^fSAD^ (magenta box in **Figure 3B**). On a whole lung level, this transition occurs on average from GOLD stages 2 to 4. By quantifying the amount and arrangement of normal and fSAD parenchyma, one can assess the severity of COPD. As fSAD is an intermediate between healthy lung and emphysema, increasing levels of emphysema have a direct effect on V and χ of fSAD. This is observed in **Figure 5** and **Figure 2**, where increasing values of V^Emph^ resulted in a drop in V^fSAD^ and increase in χ^fSAD^. These trends were reflected in our multivariable model for V^Emph^ as well (**Table 3**).

In a seminal study, McDonough and colleagues^7^ provided pathological evidence demonstrating the role of SAD in COPD progression. Using high resolution (∼10 μm) microCT to analyze frozen lung samples from lung transplant recipients with end-stage COPD, they found that widespread narrowing and destruction of the smaller airways (i.e., SAD) occurred before emphysematous lesions became large enough to be visible on standard CT imaging. They concluded that SAD might serve as an emphysema precursor. Based on their observation, we postulated that the transition observed between χ^Norm^ and χ^fSAD^ (**Figures 4** and **5**) should be observed for χ^fSAD^ and χ^Emph^. Using mean values of χ over the lungs, χ^Emph^ was found to be relatively stable, generating positive values across GOLD (**Table 4**), as well as demonstrating a weak correlation to χ^fSAD^ (ρ = -0.155, p < 0.001). Nevertheless, evaluating χ^fSAD^ and χ^Emph^ at the local level, we observe a strong association between these two readouts (**Figure 2**), which may be linked to the structural changes in the terminal airways observed using microCT of lung explants.

In a recent study, Bhatt and colleagues evaluated a CT readout, referred to as the mean Jacobian determinant of normal voxels, at varying distances from emphysematous tissue^25^. When measured at 2mm from CT voxels designated emphysema (i.e., voxel HU <-950HU), this CT-based readout was found to be predictive of spirometric decline. Our spatial analysis of a single case clearly demonstrates a transition in topologies of PRM^fSAD^ and PRM^Emph^, 1.8 cm and 1.2cm for V and χ, respectively (**Figure 2**). It is the association of topologies between PRM^fSAD^ and PRM^Emph^ at the local level that allows our machine learning model to predict spirometric decline, with an accuracy of 70%, in the absence of any emphysema readout as an input (**Figure 6**). Although the readouts reported by Bhatt and colleagues lacked quantification of SAD, there is clear agreement that lung tissue along the periphery of emphysematous tissue provides potential insight into COPD progression. Using only topologies of PRM^Norm^ and PRM^fSAD^, our patch-based ML model outperformed the whole-lung logistic regression model (**Figure 8B**). This result highlights the importance of the spatial relationship of χ^fSAD^ to χ^Emph^ to predict spirometric decline (**Figures 6** and **8**).

We acknowledge several notable limitations. COPDGene comprises over 20 study sites, making scanner variation and reconstruction kernel inconsistency inevitable. Sensitivity of PRM to scanner variability was addressed previously^26^ and although effort was made to apply PRM only to soft kernels, variability in scanner type was unavoidable. However, we included scanner vendor in our multivariable regressions and found that it did not significantly confound models. Another limitation is variation in levels of inspiration and expiration during CT acquisition. Earlier work demonstrated that even small perturbations from functional residual capacity (FRC) have an observable effect on threshold-based techniques such as PRM^26^. To limit this, we implemented QC that excluded participants based on erroneous volume changes or strong discordance with correlation between PRM^Norm^ and FEV_1_% predicted. In summary, we have demonstrated that topological features, V and χ, are able to enhance the sensitivity of PRM classifications, notably Norm and fSAD, to extent of emphysema and COPD severity. These data support the concept that as pockets of small airways disease coalesce, surrounding normal tissue is lost. Pockets of fSAD are seen to correlate with increasing presence of emphysema, independent of the amount of fSAD present. We further demonstrated that local levels of χ^fSAD^ and χ^Emph^ correlate, which may be explained by bronchiolitis along the periphery of emphysematous tissue observed by McDonough and colleagues using microCT. In addition, we demonstrated that local values of V and χ for PRM^Norm^ and PRM^fSAD^ provide sufficient information to predict spirometric decline, even in the absence of any prior knowledge of emphysema. Our study provides a unique strategy to detect subtle changes in lung parenchyma that may progress to emphysema. This approach to monitoring extent and arrangement of Norm and fSAD offers insight into COPD phenotypes and provides improved prognostic information that has relevance in clinical care and future clinical trials.

## Data Availability

The datasets presented in this study are not readily available because they are part of an NIH sponsored clinical trial and require a data use agreement to be signed. For access to COPDGene data visit https://www.copdgene.org/phase-1-study-documents.htm for instructions.

## Conflicts of Interest and Sources of Funding

### Conflicts of Interest

Wassim W. Labaki reports personal fees from Continuing Education Alliance. Benjamin A. Hoff and Craig J. Galban are co-inventors and patent holders of tPRM, which the University of Michigan has licensed to Imbio, LLC. Craig J. Galban is co-inventor and patent holder of PRM, which the University of Michigan has licensed to Imbio, LLC. Benjamin A. Hoff and Craig J. Galban have financial interest in Imbio, LLC. Charles R. Hatt is employed by Imbio, LLC. David A. Lynch reports funds paid to the institution from NIH and personal payments from Boehringer Ingelheim. MeiLan K. Han reports personal fees from GlaxoSmithKline, AstraZeneca, Boehringer Ingelheim, Cipla, Chiesi, Novartis, Pulmonx, Teva, Verona, Merck, Mylan, Sanofi, DevPro, Aerogen, Polarian, Regeneron, Amgen, UpToDate, Altesa Biopharma, Medscape, NACE, MDBriefcase and Integrity. She has received either in kind research support or funds paid to the institution from the NIH, Novartis, Sunovion, Nuvaira, Sanofi, AstraZeneca, Boehringer Ingelheim, Gala Therapeutics, Biodesix, the COPD Foundation and the American Lung Association. She has participated in Data Safety Monitoring Boards for Novartis and Medtronic with funds paid to the institution. She has received stock options from Meissa Vaccines and Altesa Biopharma. For the remaining authors none were declared.

### Funding

This work was supported by the National Heart, Lung, and Blood Institute of the National Institutes of Health Grants R01 HL139690 (to Craig J. Galban), R01 HL150023 (to MeiLan K. Han, Craig J. Galban, and Charles R. Hatt) and T32 HL 007749 (to Jennifer M. Wang) and by the National Heart, Lung, and Blood Institute of the National Institutes of Health Grants U01 HL089897 and U01 HL089856, and NIH contract 75N92023D00011, which support the COPDGene study. The COPDGene study (NCT00608764) is also supported by the COPD Foundation through contributions made to an Industry Advisory Committee that has included AstraZeneca, Bayer Pharmaceuticals, Boehringer-Ingelheim, Genentech, GlaxoSmithKline, Novartis, Pfizer and Sunovion.

## Acknowledgements

We acknowledge the COPDGene investigators for their role in the study providing data for this project:

**Administrative Center**: James D. Crapo, MD (PI); Edwin K. Silverman, MD, PhD (PI); Barry J. Make, MD; Elizabeth A. Regan, MD, PhD

**Genetic Analysis Center**: Terri Beaty, PhD; Ferdouse Begum, PhD; Peter J. Castaldi, MD, MSc; Michael Cho, MD; Dawn L. DeMeo, MD, MPH; Adel R. Boueiz, MD; Marilyn G. Foreman, MD, MS; Eitan Halper-Stromberg; Lystra P. Hayden, MD, MMSc; Craig P. Hersh, MD, MPH; Jacqueline Hetmanski, MS, MPH; Brian D. Hobbs, MD; John E. Hokanson, MPH, PhD; Nan Laird, PhD; Christoph Lange, PhD; Sharon M. Lutz, PhD; Merry-Lynn McDonald, PhD; Margaret M. Parker, PhD; Dmitry Prokopenko, Ph.D; Dandi Qiao, PhD; Elizabeth A. Regan, MD, PhD; Phuwanat Sakornsakolpat, MD; Edwin K. Silverman, MD, PhD; Emily S. Wan, MD; Sungho Won, PhD

**Imaging Center**: Juan Pablo Centeno; Jean-Paul Charbonnier, PhD; Harvey O. Coxson, PhD; Craig J. Galban, PhD; MeiLan K. Han, MD, MS; Eric A. Hoffman, Stephen Humphries, PhD; Francine L. Jacobson, MD, MPH; Philip F. Judy, PhD; Ella A. Kazerooni, MD; Alex Kluiber; David A. Lynch, MB; Pietro Nardelli, PhD; John D. Newell, Jr., MD; Aleena Notary; Andrea Oh, MD; Elizabeth A. Regan, MD, PhD; James C. Ross, PhD; Raul San Jose Estepar, PhD; Joyce Schroeder, MD; Jered Sieren; Berend C. Stoel, PhD; Juerg Tschirren, PhD; Edwin Van Beek, MD, PhD; Bram van Ginneken, PhD; Eva van Rikxoort, PhD; Gonzalo Vegas Sanchez-Ferrero, PhD; Lucas Veitel; George R. Washko, MD; Carla G. Wilson, MS; PFT QA Center, Salt Lake City, UT: Robert Jensen, PhD

**Data Coordinating Center and Biostatistics, National Jewish Health, Denver, CO**: Douglas Everett, PhD; Jim Crooks, PhD; Katherine Pratte, PhD; Matt Strand, PhD; Carla G. Wilson, MS

**Epidemiology Core, University of Colorado Anschutz Medical Campus, Aurora, CO**: John E. Hokanson, MPH, PhD; Gregory Kinney, MPH, PhD; Sharon M. Lutz, PhD; Kendra A. Young, PhD

**Mortality Adjudication Core**: Surya P. Bhatt, MD; Jessica Bon, MD; Alejandro A. Diaz, MD, MPH; MeiLan K. Han, MD, MS; Barry Make, MD; Susan Murray, ScD; Elizabeth Regan, MD; Xavier Soler, MD; Carla G. Wilson, MS

**Biomarker Core**: Russell P. Bowler, MD, PhD; Katerina Kechris, PhD; Farnoush Banaei-Kashani, Ph.D We would also like to acknowledge our copy editor Lee Olsen for her assistance in preparing this manuscript.

## Appendices

## Appendix 1: Quality Control (QC) Protocol

QC was performed in two steps (e.g., see 3 and 4 in exclusion diagram below), using GOLD grade, segmented lung volume change (ΔV = inspiration volume – expiration volume, as a function of segmented voxels), and a correlation test metric (Q) defined as the absolute value of the difference between standard scores of FEV_1_% predicted and %PRM^Norm^, reported to be highly correlated in COPD studies.^8^

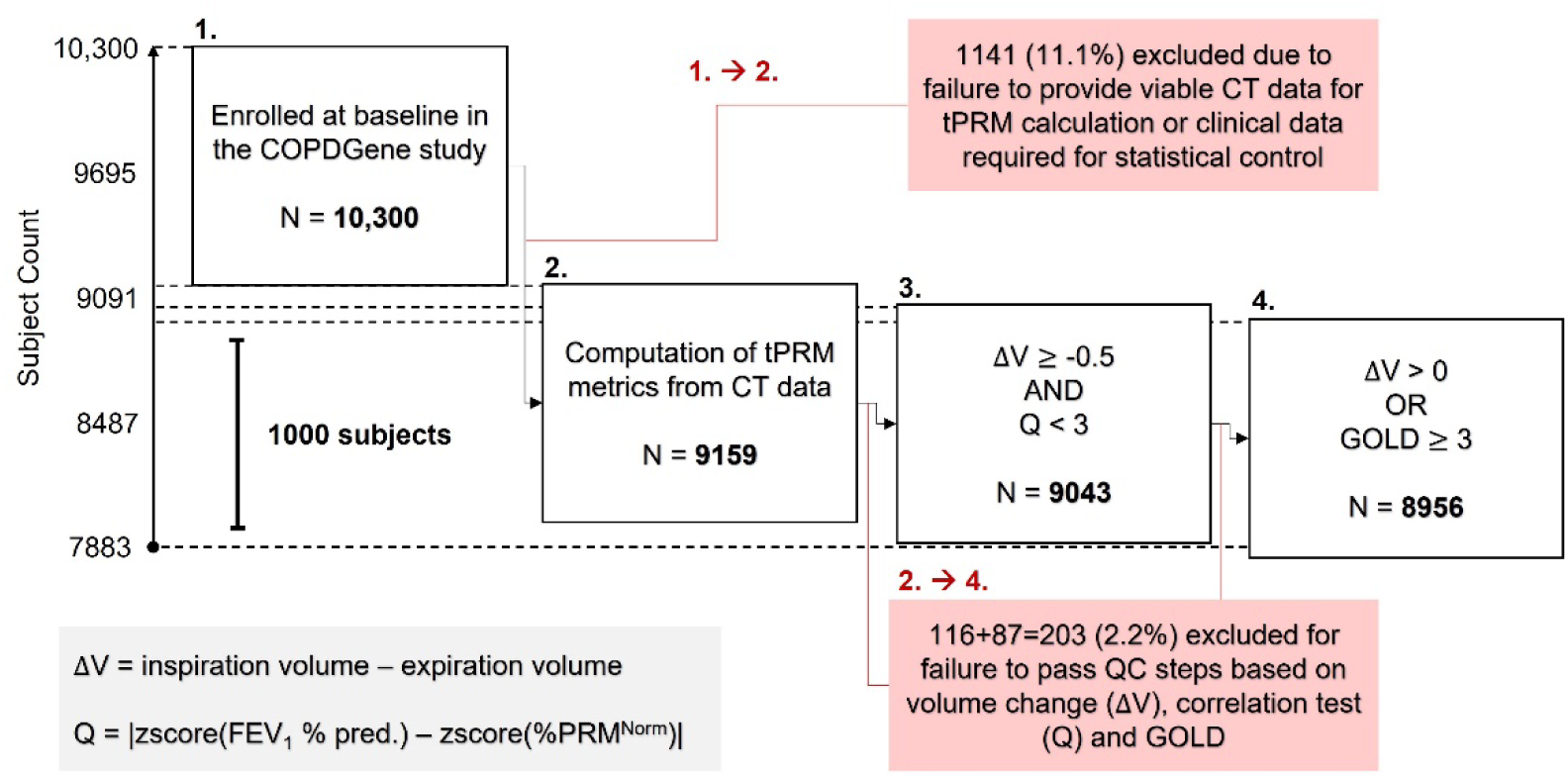

Specifically, imaging QC tests for exclusion were applied consecutively as follows:

### 1. ΔV < -0.5 L OR Q ≥ 3

A large negative volume change, here defined as greater than 0.5 L, often indicates transposition of intended respiratory stages (expiration/inspiration), due to faulty maneuver or data handling error. In addition, we test here if a deviation of equal to or greater than 3 standard deviations from the expected positive correlation between FEV_1_% predicted and %PRM^Norm^ has occurred.

### 2. ΔV ≤ 0 L and GOLD < 3

This second step goes on to test if a non-severe COPD participant (GOLD < 3) has zero or negative volume change. N.B. here and in step 1 we have considered that there may be participants with severe disease that have some abnormal volume changes (close to 0 due to very limited lung function).

## Appendix 2: Dictionary Learning Algorithm

Image sparsity has emerged as a significant property of images and sparsity-based regularization has been used for various image processing applications.^18–20, 24, 27–30^ Sparse image representations are at the heart of many modern approaches to medical image classification and include ^31–33^. The sparse model assumes that each patch within an image can be accurately represented using a few elements of a basis set called a dictionary. For image classification problems a separate class-specific dictionary is learnt from patches belonging to each class of images. In this work, we have developed a multiview task-driven dictionary learning algorithm – a novel approach that aims to learn discriminative dictionaries for each class from multiple views of the data in a joint fashion by imposing group sparsity constraints^34^.

### Dictionary Learning

Our proposed method utilizes an overcomplete dictionary *D* constructed from the CT images, which is an *n* × *K* matrix whose columns represent *K* “atoms” of size *n*, where an “atom” is a sparse coefficient vector (i.e., a vector of weights/coefficients in the sparse basis). We train a separate dictionary for each class. Each dictionary *D*_*i*_ represents the image patches from class *i* reasonably well, but at the same time represents the image patches from the other classes quite poorly. There are several ways to train/learn a dictionary.^35^ In this work, we have adopted the task-driven dictionary learning algorithm proposed by Mairal et al.^36^ To train them, we solve a combinatorial optimization problem, where an approximate solution is obtained by alternating between a greedy sparse coding step using the current dictionary estimate, and a dictionary update step.

### Sparse Coding

We assume that any image patch *x* in a CT image can be represented as a sparse linear combination of the atoms of the dictionary *D* as: *x* ≈*Dα*, where *D* is the sparse coefficient vector. Given a dictionary, *D*, the goal in sparse coding is to find a sparse coefficient vector *D*. This requires solving a second optimization problem, the optimal solution to which is found using a greedy approach such as an orthogonal matching pursuit algorithm.^37^

### Classification

In sparse representation-based classification, an image patch *x* is classified according to how well the patch is represented by the class-specific dictionaries. Once a dictionary *D_i_* has been trained for each class *i*, classification of a new image patch *x*_new_ is performed by evaluating the reconstruction/representation errors for different classes. From the class representation errors a pseudo-probability measure *P_i_* is computed and the image patch is assigned to the class that has the maximum probability value

### Training

The dictionary learning model was trained on a desktop workstation running a 64-bit Windows operating system (Windows 10) with an Intel Xeon W-2123 CPU at 3.6GHz with 128GB DDR4 RAM. The x-, y-, and z-dimensions of each image in our dataset were x = 512, y = 512, and z ∼ 1250. CT lung scans from N = 4483 cases from the COPDGene phase 1 dataset who had follow up examination were considered. A representative 2D slice of a donor lung CT image is shown in **Figure 3A** of the manuscript. The lungs within these CT images were then automatically segmented using in-house software developed using MATLAB R2020a (MathWorks, Natick, MA). Our dataset for this study consists of a total N = 4483 lung CT images belonging to two categories: i) N = 1516 CT lungs that were described as fast progressors with a change of FEV_1_ ≥ -60ml/yr, referred to as class 1, and ii) N = 2967 CT lungs that were described as slow progressors with a change of < -60ml/yr in their FEV_1_, referred to as class 2. We used 35% of the data for training and the remaining 65% for testing. A total of 8,000,000 2D image patches from the three (axial, coronal, and sagittal) views from each of the prior maps (tPRM maps V^Norm^, V^fSAD^, χ^Norm^, and χ^fSAD^) were extracted from the training data for each class to train the dictionaries. The proposed dictionary learning algorithm was developed using MATLAB R2020a software (MathWorks, Natick, MA). We used the sparse modeling software (SPAMS) toolbox^38^ for the orthogonal matching pursuit optimization algorithm to efficiently optimize the dictionary elements. The hyper parameters of the dictionary learning algorithm include the image patch size *l*, the number of dictionary bases *K* for each dictionary, the sparsity controlling parameter *λ*, and the positive regularization parameter *ρ* in sparse coding. The optimal values for these parameters were automatically selected on a validation set (randomly chosen from within the training data) using the receiver operating characteristic (ROC) curves, by varying one parameter at a time while keeping the others fixed and choosing that value of the parameter that maximizes the area under the curve (AUC) of the ROC curve. The parameters of the dictionary learning algorithm were set to *l* = 25, *K* = 512, *λ* = 0.0015, and *ρ* = 0.006.

## Notes

### Author Declarations

Our study was a secondary analysis of data from COPDGene (ClinicalTrials.gov: NCT00608764). The institutional review boards of all 21 centers participating in the COPDGene study gave ethical approval for the work of that study.

### Summary of Updates

All supplemental material has been incorporated into the main text. This change provides a more accessible and comprehensive picture of our dictionary learning model; specifically, figures 6 and 7 are discussed in sequence to demonstrate how the model can identify COPD patients as fast or slow progressors. We have also added an author, Dr. Ravi Pal.

